# Prediction of evolution of the second wave of Covid-19 pandemic in Italy

**DOI:** 10.1101/2020.11.24.20238139

**Authors:** Ignazio Ciufolini, Antonio Paolozzi

## Abstract

A relevant problem in the study of the Covid-19 pandemic is the study of its temporal evolution. Such evolution depends on a number of factors, among which the average rate of contacts between susceptible and infected individuals, the duration of infectiousness and the transmissibility, that is the probability of infection after a contact between susceptible and infected individuals. In a previous study, we analyzed the potentiality of a number of distributions to describe the evolution of the pandemic and the potentiality of each distribution to mathematically predict the evolution of the pandemic in Italy. Since the number of daily tests was changing and increasing with time, we used the ratio of the new daily cases per swab. We considered distributions of the type of Gauss (normal), Gamma, Beta, Weibull, Lognormal and in addition of the type of the Planck blackbody radiation law. The Planck law, describing the amount of energy of the electromagnetic radiation emitted by a black body at each wavelength or at each frequency, marked in 1900 the beginning of Quantum Mechanics. The result of our analysis was that, among the considered distributions, the Planck law has the best potentiality to mathematically predict the evolution of the pandemic and the best fitting capability. In this paper, we analyze the time evolution of the second wave of the Covid-19 pandemic in Italy and in particular we predict the ratio of the new daily cases per swab at Christmas 2020 using the data in the interval from 17 Oct to 21 Nov. According to Figure 4 and Figure 8, the prediction for such a ratio around Christmas is approximately within 6% and 7%. In this study there is also an attempt to account for the effects of the governmental containment measures.

## Section 1. Introduction

Planck’s law is one of the outstanding laws of physics, it describes the energy of the radiation emitted by a black body at temperature T as a function of the frequency. Derived in 1900 by Max Planck^1^, with the idea of energy quantization, marked the beginning of Quantum Mechanics. It is impressive that the electromagnetic radiation emitted about 400,000 years after the Big Bang is today observed as Cosmic Microwave Background (CMB) radiation, that is the cooled remnant of the Big Bang radiation, and is well described as blackbody radiation at 2.728 K (Fig. 1A). Furthermore, Wien law, a consequence of the Planck law, is used to determine the temperature of a star by measuring the peak frequency of the electromagnetic radiation emitted by the star. The Planck law expressed as number of photons per unit area, unit time and unit frequency, ν, emitted by a black body at thermal equilibrium with temperature T, is: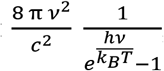, where *h, c* and *k*_*B*_ are respectively Planck constant, speed of light and Boltzmann constant^1^. In a previous paper^2^, we analyzed the evolution of the pandemic in Italy using various distributions of the type of the Gauss (normal), Gamma, Beta, Weibull and Lognormal distributions and of the type of the Planck blackbody law (with two or three parameters). Since the number of daily swabs was changing and increasing with time, we analyzed the experimental data consisting in the number of new daily cases per daily swabs. It turned out that the best distribution to both fit the experimental data of the pandemic and to predict its future evolution in Italy is a Planck distribution. In this paper we analyze the evolution of this second wave of the pandemic in Italy, using the Planck distribution with three parameters to make predictions on the number of new daily cases per swab in particular during the next Christmas holidays.

**Fig. 1.**
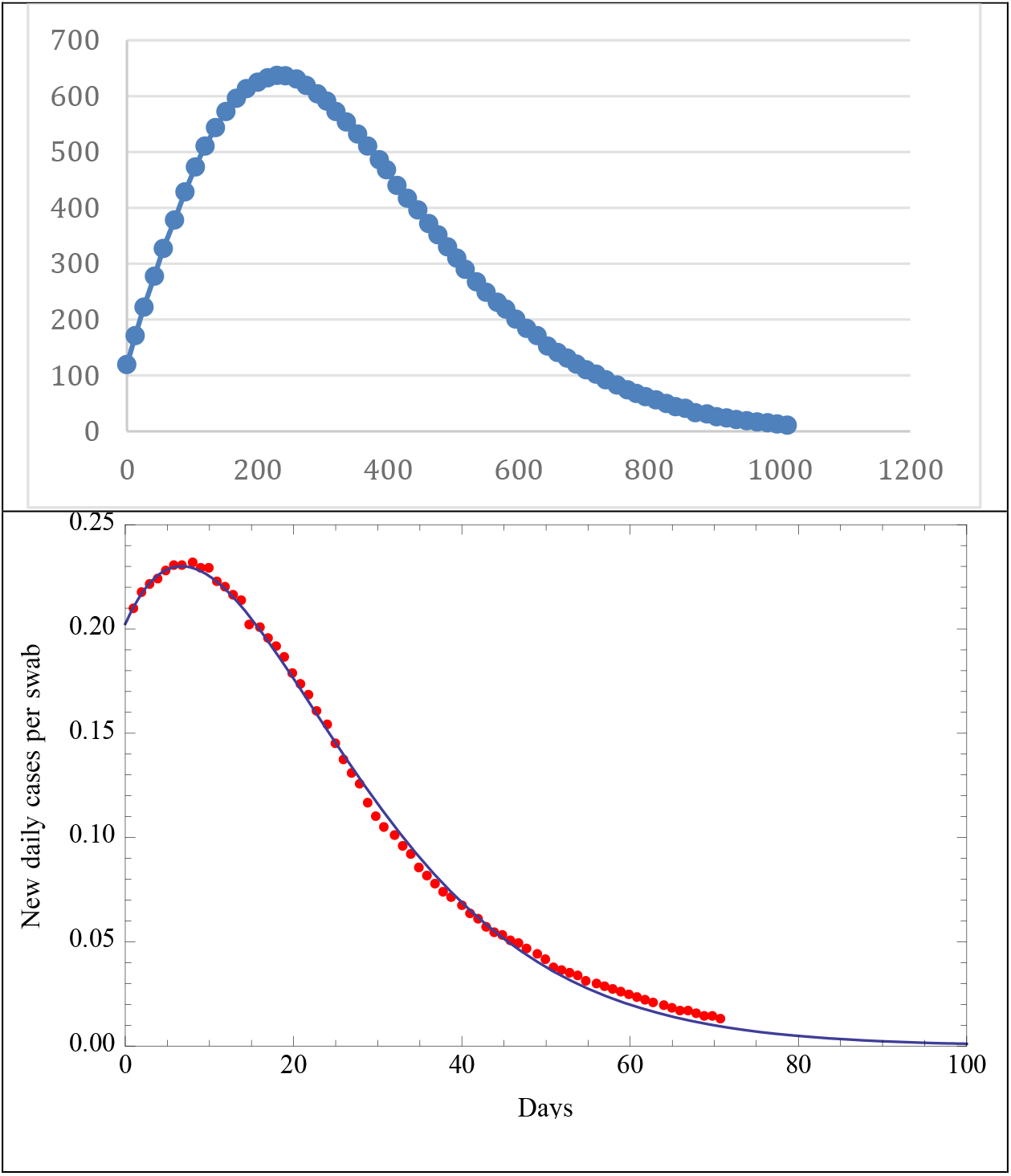
**A**. The Cosmic Background Microwave Radiation fitted with a Planck distribution4 at 2.728 K. **B**. First wave of Covid-19, moving average over four weeks of the ratios of daily cases per swab for Italy and its best fit curve using a Planck’s distribution with three parameters. The red dots are the moving average of the ratios of new daily cases per swab and the solid black line is the best fit Planck’s distribution.

## Section 2. Planck law and the evolution of the pandemic

To describe the evolution of the pandemic, we analyzed the evolution of the new daily cases per daily swab using a Planck distribution with three parameters. The parameters of the Plank’s law can take into account some of the parameter considered for instance in the SIR model of the pandemic3 such as the daily number of infected persons, the transmissibility, that is the probability of infection due to the contact between a susceptible and infected individual, the average rate of contacts between susceptible and infected individuals (more persons the infected individual will contact, more persons will be infected) and the duration of infectiousness (more will last the infection of a contagious individual, more persons will be infected). In Fig. 1B, one can observe the impressive potentiality of the Planck’s distribution to fit the experimental data of the pandemic, we first took the moving average of over four weeks of the ratios of daily cases per swab of Italy and we then fitted it with the Planck distribution Eq. (1).

In a previous paper^2^, using a Planck’s distribution with three parameters, we fitted the experimental data of new daily cases per swab in Italy and we predicted, a few weeks ahead of time, that such ratio would decrease below 0.0043 on May 27 with a three-sigma uncertainty of +/- 9 days calculated using a Monte Carlo simulation with 25000 runs. Indeed, in agreement with our mathematical estimate, the first day in which such low ratio was below 0.0043 was June 4 when the ratio reached the value of 0.0035 (its historical minimum in Italy from February 25 to June 4). We also attempted to estimate the uncertainty of our prediction, possibly taking into account the various systematic errors, by calculating the spread of the pandemic among the twenty Italian regions and we obtained a one-sigma uncertainty of about 11.4 days. In Fig. 2 we reproduce our fit and mathematical prediction^2^ for the number of new daily cases per swab, obtained on May 16. It agrees with a relative error of only 39% with the number of new daily cases per swab measured in June 6, i.e., three weeks later.

**Fig. 2.**
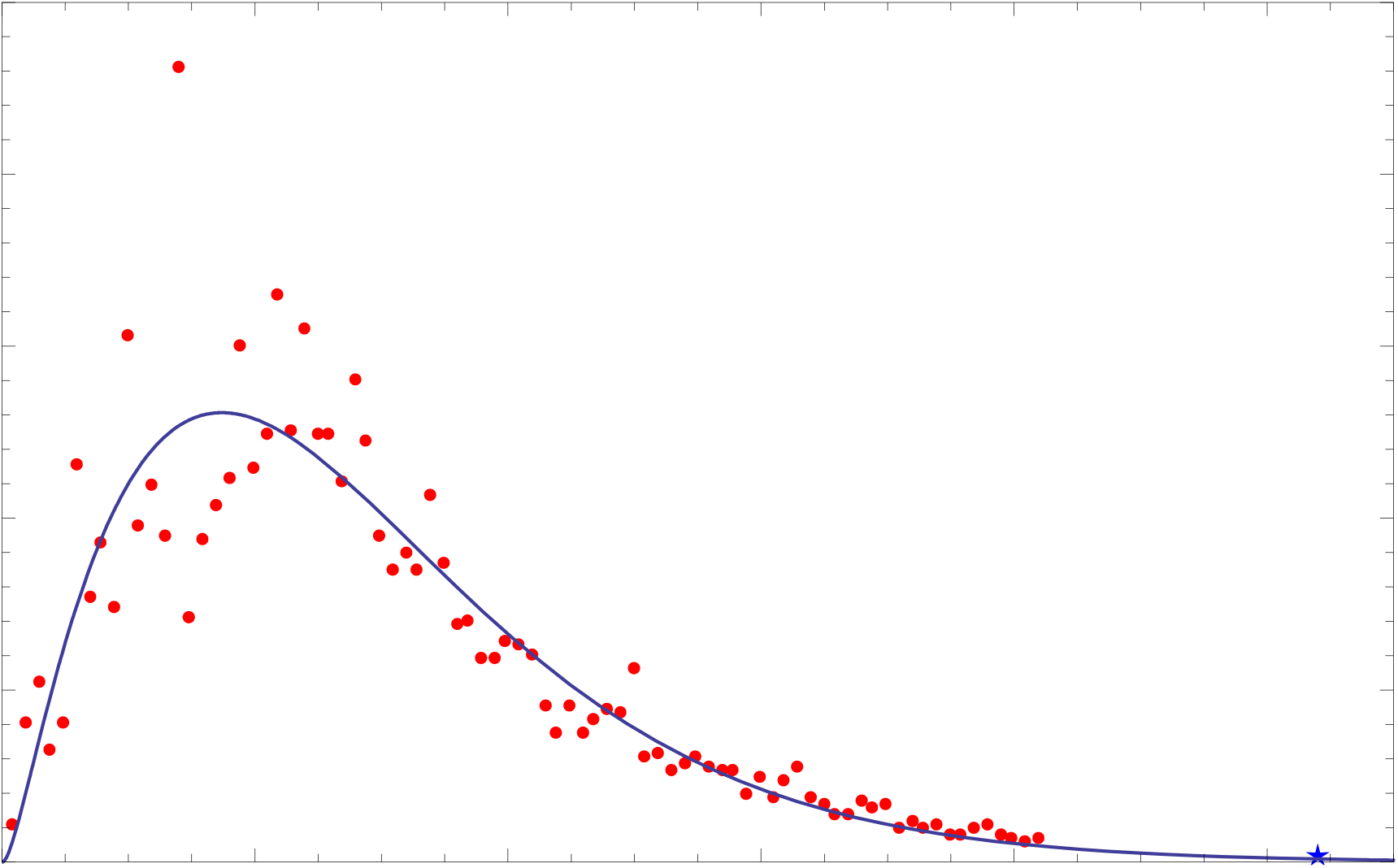
The red dots are the new daily cases per swab in Italy from February 25 to May 16, the black solid line is the best fit Planck’s function which fits these cases per swab from February 25 to May 16, it is extrapolated up to June 13. The blue star represents the real value of cases per swab measured on June 6. The difference between such measured value and that extrapolated from the Planck’s fitting function is about 39%.

In Fig. 3 the daily cases-to-swab ratios smoothed with 7-days moving average are reported. The beginning of the curve corresponds to the first day in which data on nasopharyngeal swabs are available i.e. the 25 February 2020 and the last point corresponds to the 22 November 2020. The two waves of the pandemic diffusion in Italy are clearly visible.

**Fig. 3.**
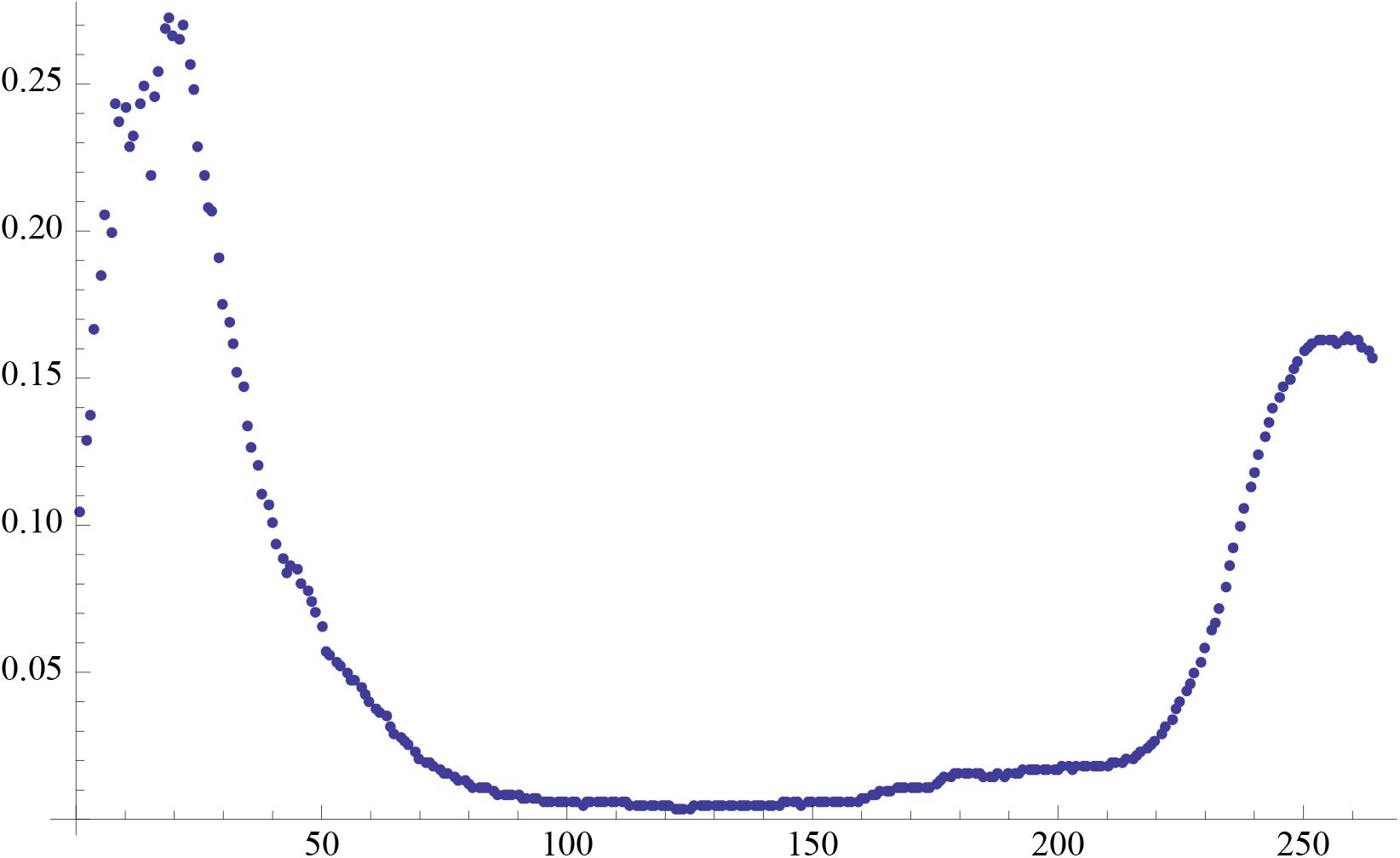
Behavior of the pandemic in Italy from the 25 February 2020 to 22 November 2020. The vertical axis reports the positive daily cases-to-swab ratios smoothed with a 7-days moving average. The horizontal axis repots the days.

## Section 3. Second wave of Covid-19 in Italy

We now show the results of this second wave of Covid-19 analyzed using the Planck’s law. In Fig. 4 are reported the daily cases-to-swab ratios for 36 days starting from 17 October 2020, The continuous curve is the best fit of the data using a three parameter Planck’s law. Christmas is indicated by the vertical blue line.

**Fig. 4.**
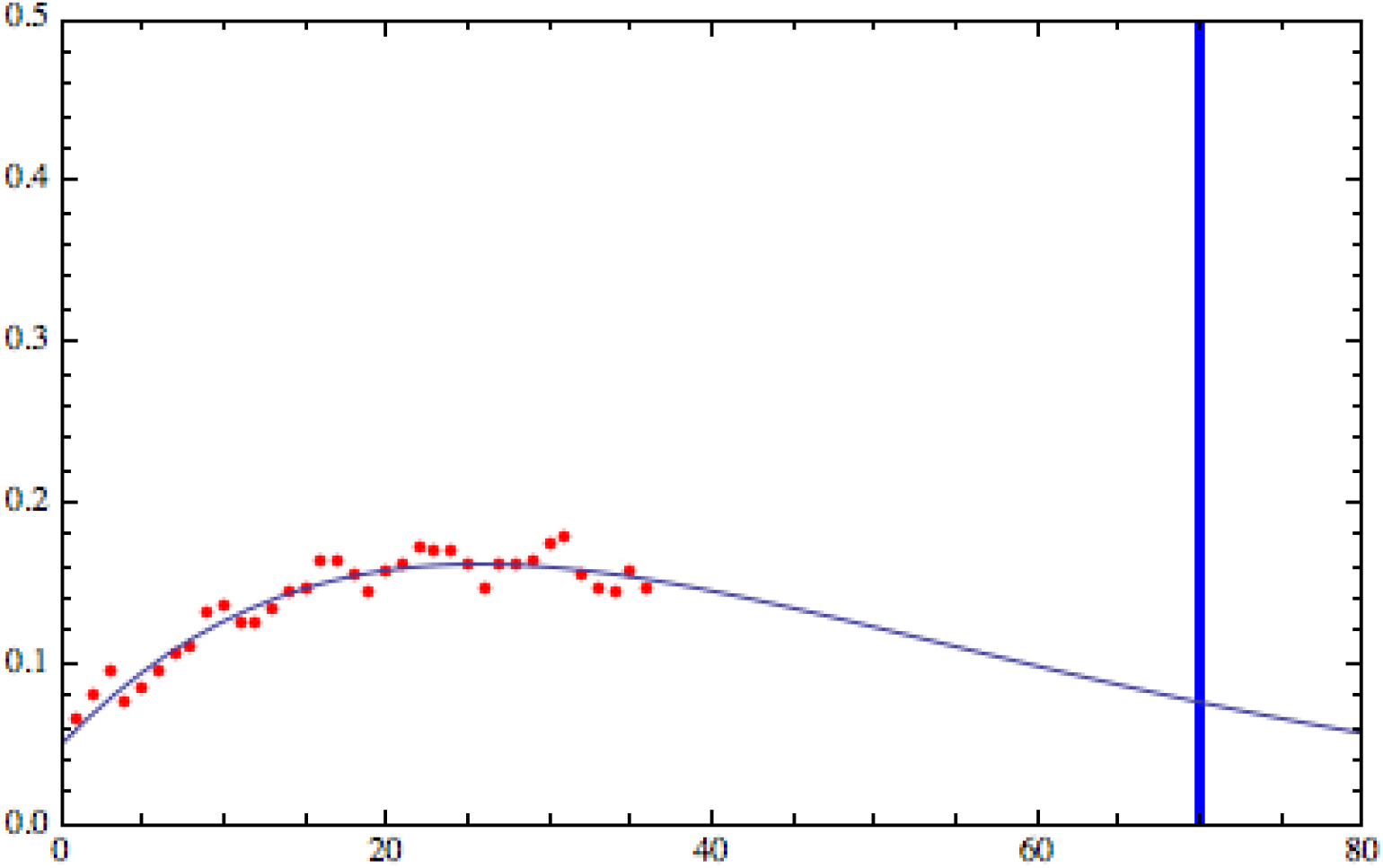
Positive daily cases-to-swab ratios along with the Planck’s law fitting function as a function of days. Data points are 36 starting from 17 October 2020. The value of this ratio at Christmas is 0.076 i.e. about 7%.

In spite of the fact that the fluctuations of the data are very limited, those can be reduced using the moving average technique. In Fig. 5 are reported the 7-days moving average ratios of this second wave of the pandemic.

**Fig. 5.**
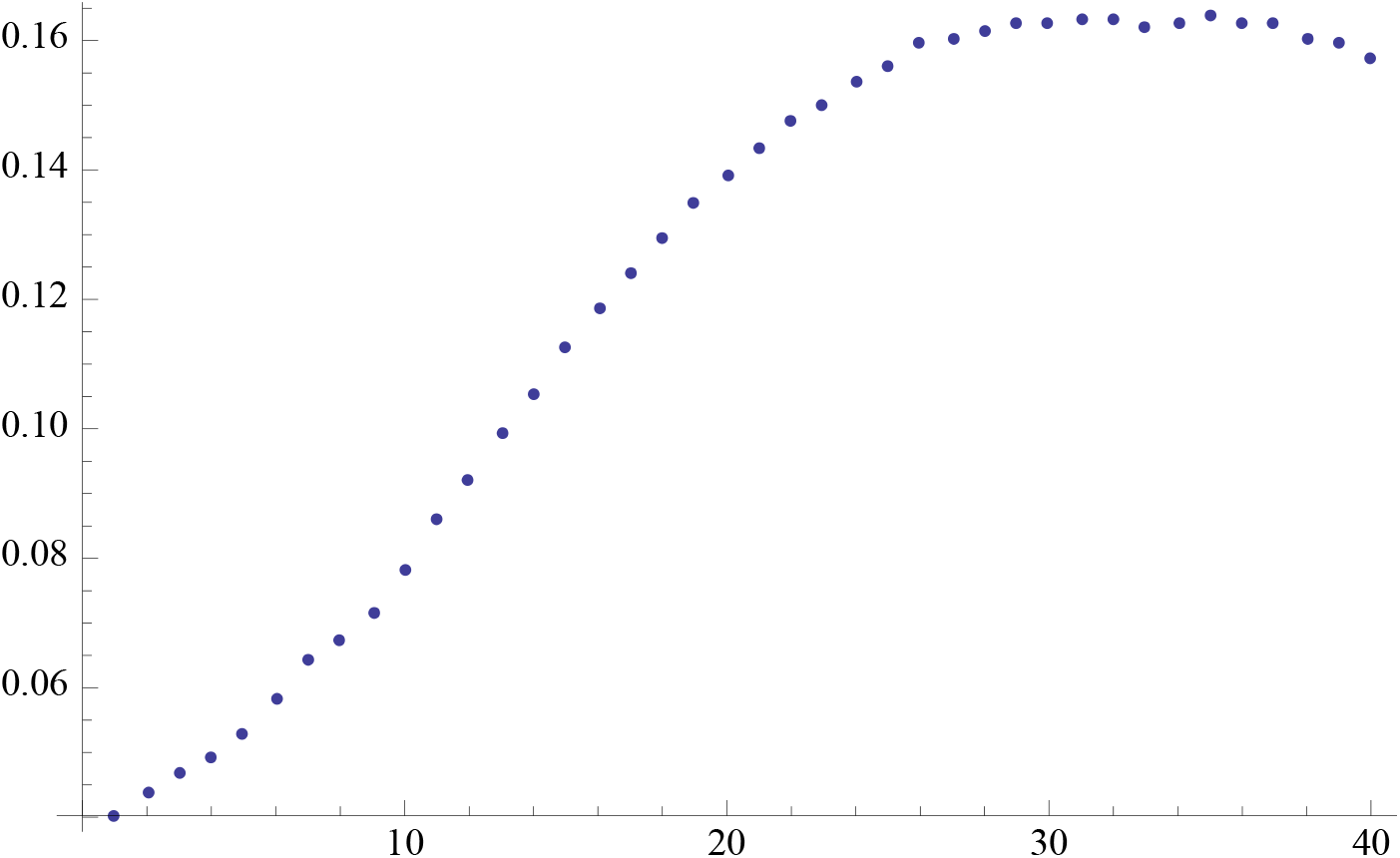
Second wave Covid-19 pandemic in Italy reported as 7-days moving average. Vertical axis reports positive daily cases-to-swab ratios. Horizontal axis reports days.

In Figs. 6 and 7 are reported the 3-days and 7 days moving average ratios with the corresponding fit with the Planck’s law for 34 and 30 points respectively. The fluctuations reduce as expected showing an almost perfect match in the case of 7 days moving average case.

**Fig. 6.**
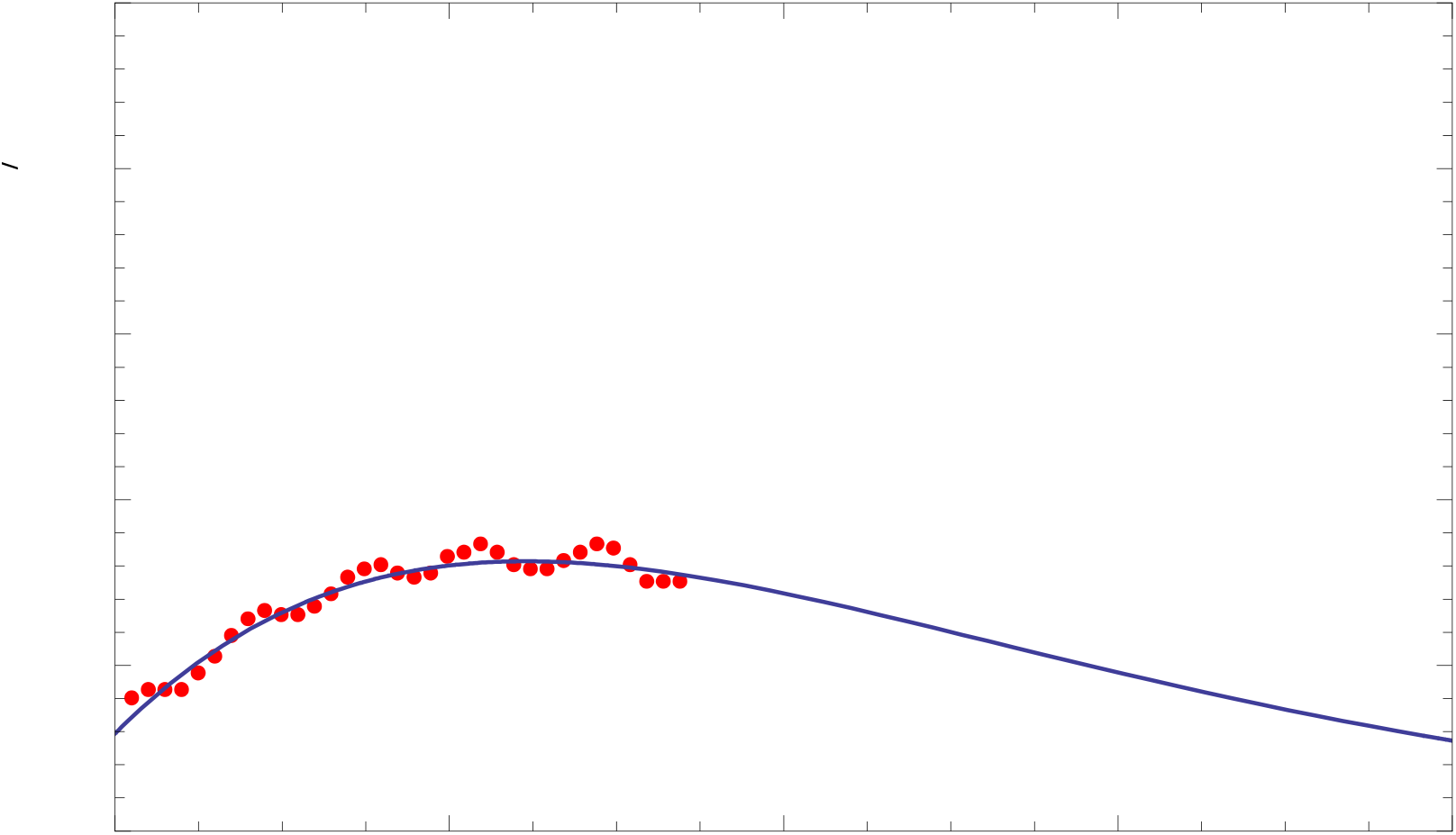
Three-days moving average of positive daily cases-to-swab ratios along with the Planck’s law fitting function vs days. Data points are 34. Horizontal axis reports the days.

**Fig. 7.**
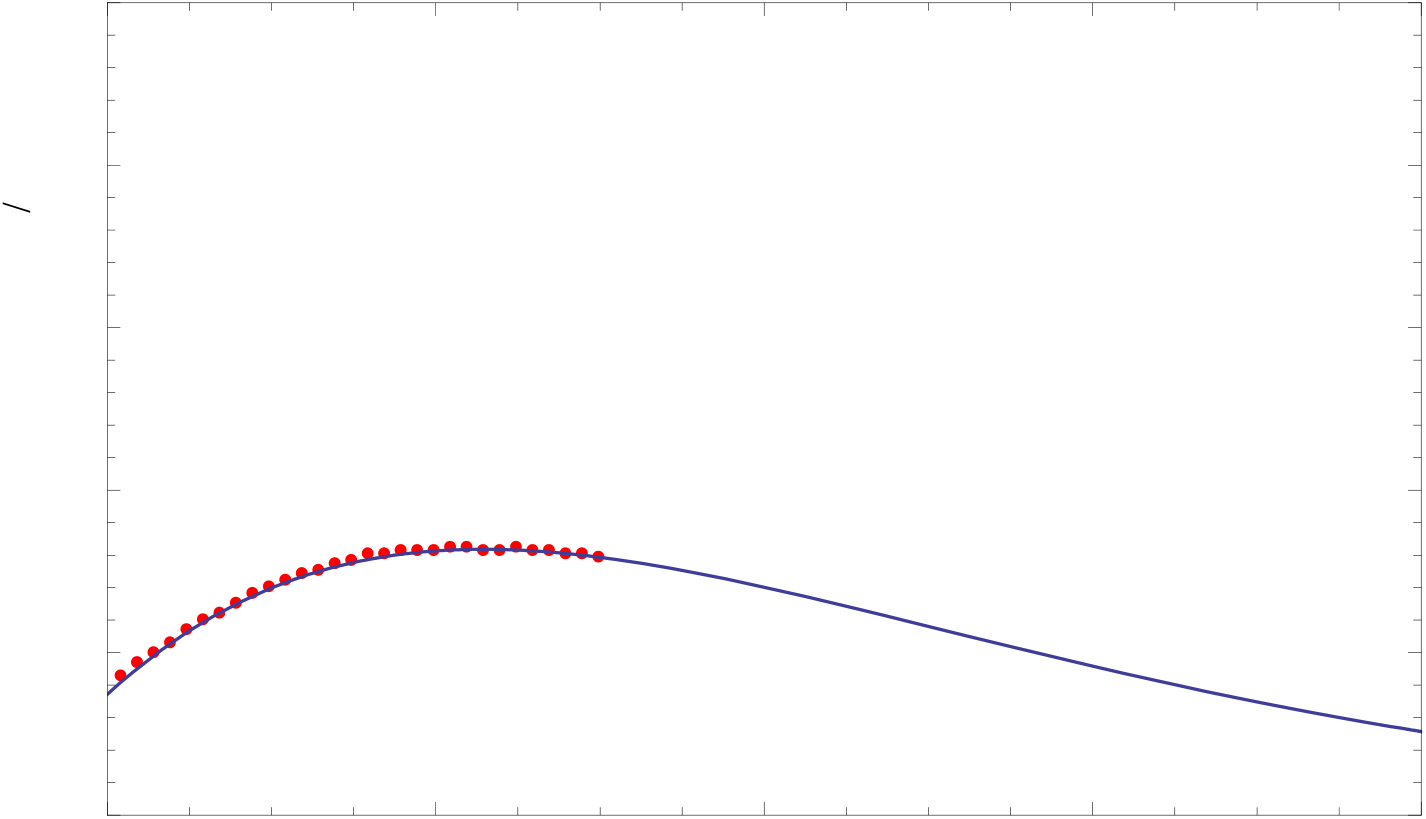
Seven-days moving average of positive daily cases-to-swab ratios along with the Planck’s law fitting function. Data points are 30. Horizontal axis reports the days.

Starting from mid-October the Italian government has enacted four decrees that gradually introduced containment measures finalized to reduce the pandemic. Specifically the four DPCM (Decreto del Presidente del Consiglio dei Ministri) were in effects on 13, 18, 24 October 2020 and 3 November 2020. Many parameters, as mentioned earlier, should be taken into account for a better prediction of the effects of the decrees on the pandemic evolution, however a simpler approach can be followed as we suggest here. We choose 30 days as a reasonably large number of days to fit the 45 days of data available. We then fit 30 days of data starting from 07 Oct, 12 Oct, 17 Oct, 22 Oct. The four values of the positive daily cases-to-swab ratios at Christmas are then compared for the different time intervals used. The results are reported in Fig. 8. We observe that the values of the ratios reduce while getting closer to the last days. This is what one expects if containment measures and individual behaviour are efficiently applied. Indeed the last days are the most significant because are affected by the containment measures. In this way we can visually estimate the asymptotic behaviour of the curve that approaches the value of about 6%. The ratio of the last point in Fig. 8 is 0.0667.

**Fig. 8.**
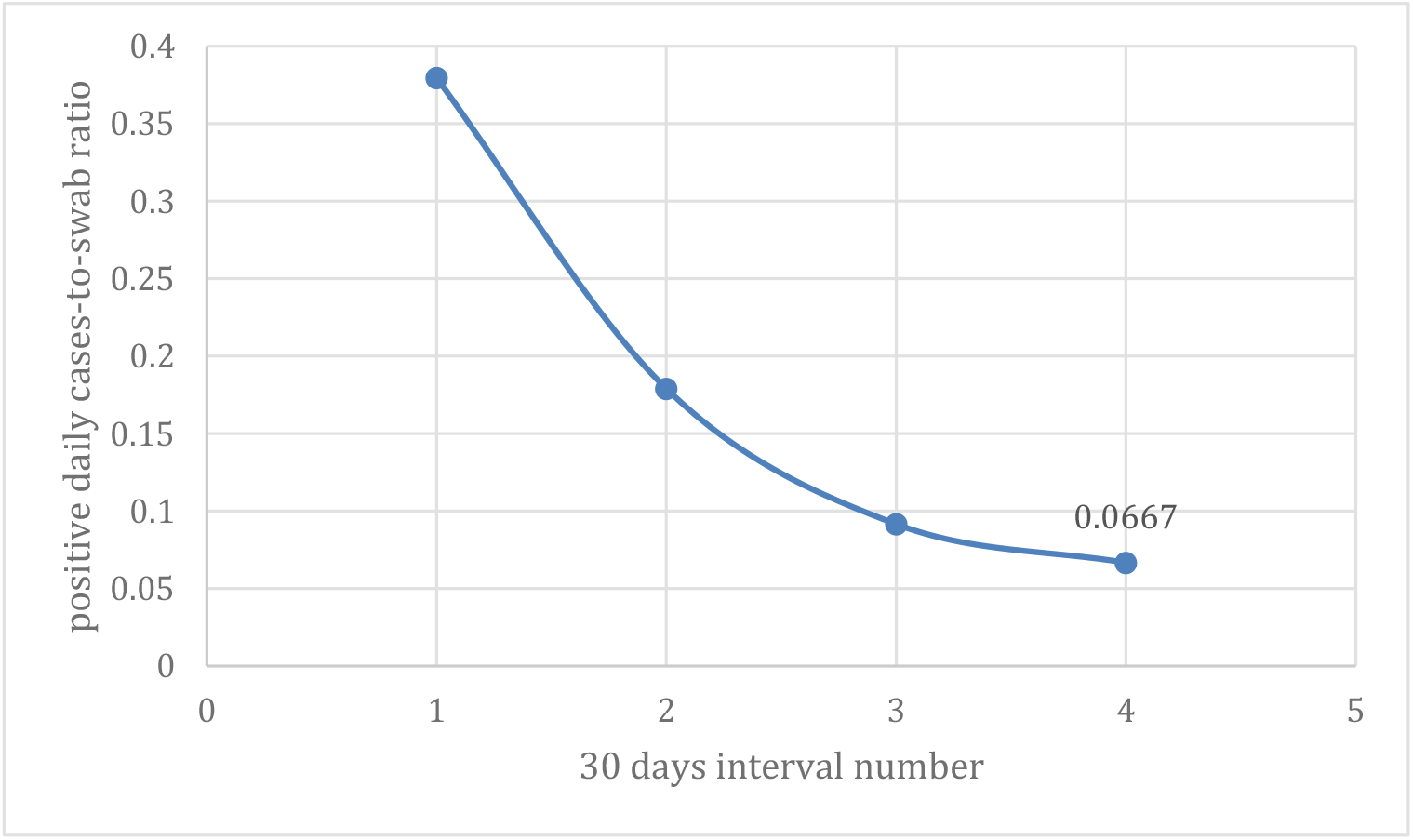
Positive daily cases-to-swab ratios at Christmas for different intervals of 30 days. The asymptotic behaviour of the curve approaches about 6%. The last point corresponds to 6.67% of the daily positive cases with respect to the number of daily swabs.

## Section 4. Conclusions

Planck’s blackbody law is one of the outstanding achievements of the physical sciences. It marked the beginning of Quantum Mechanics and remarkably described the cosmic electromagnetic radiation emitted shortly after the Big Bang observed today as cosmic microwave radiation pervading the universe. It turns out that, in addition to the various applications in physics and astronomy, it can well mathematically describe the evolution of the pandemic and well predict its future evolution, which however depends on a number of social parameters that cannot be predicted. We applied the Planck’s law to analyze the experimental data of the new daily cases per swab of Italy during this second wave of the pandemic, and showed that the Planck’s distribution confirms its remarkable good fitting capability and we believe a good potential for predicting the pandemic evolution. The parameters appearing in the Planck’s distribution to fit the pandemic are related to the relevant parameters of the pandemic such as the daily number of infected persons, the probability of infection due to contact between a susceptible and an infected individual, the average rate of contacts between susceptible and infected individuals and the duration of infectiousness. Of course any unpredictable change in these parameters would affect the mathematical prediction with the Planck’s distribution that assumes constant parameters. The time evolution of the second wave of the Covid-19 pandemic in Italy is studied. In particular the ratio of the new daily cases per swab in the coming Christmas, according to Figure 4 and Figure 8, is found to be approximately within 6% and 7%.

## Data Availability

The manuscript make use of publicly available data: the sources are referred in the text.

